# Original Investigation: Evolution of Long-Term Cardiac Tumors in Patients with Tuberous Sclerosis

**DOI:** 10.1101/2025.04.18.25326091

**Authors:** Nathalia Conci Santorio, Anna Christina Ribeiro, Nilson Bossle Conci, Gardênia da Silva Lobo Oishi, Pandreli Testa Santorio, Maria Rosa Quadrado Matos, Nana Miura Ikari, Fábio Fernandes, Viviane Tiemi Hotta

## Abstract

**Background:** The tuberous sclerosis complex is an autosomal dominant genetic disorder, resulting from mutations in the tumor suppressor genes Tuberous Sclerosis Complex 1 or 2. Cardiac rhabdomyomas are the most frequent initial manifestation and are the main causes of mortality in patients under 10 years of age. Data on the evolution and follow-up of Brazilian patients with rhabdomyomas associated with tuberous sclerosis are scarce. This study aims to describe diagnostic aspects and clinical features observed during the follow-up in a high-complexity cardiological institution.

**Methods:** It is a retrospective, descriptive, single-center study based on the collection of data from the institution’s medical records. Patients in the pediatric age group (zero to 18 years) and adult age group (over 18 years) of both genders were included, with a confirmed diagnosis of tuberous sclerosis based on the updated 2021 criteria, from January 1997 to January 2024. Patients with at least two serial transthoracic echocardiograms performed at the service were included, and patients with doubtful diagnoses and incomplete records were excluded.

**Results:** Among the 69 patients evaluated, 42 (60.86%) had cardiac tumors, with 41 rhabdomyomas and one pericardial lipoma, with a mean follow-up time of 6 years. Cardiac tumors were more frequently multiple, in 75.6% of cases. The vast majority of patients with rhabdomyomas were asymptomatic in both evaluations (73.8% and 85.71%, respectively); however, episodes of arrhythmia were recorded in 21.43% of the sample during follow-up. Only one patient presented with ventricular dysfunction, and one patient required surgical treatment, resulting in death. Regarding clinical evolution, the most frequent presentation was incomplete involution of the mass in 76.2% of cases, with complete regression in 16.7% of cases and maintenance, increase, or need for surgical treatment in 7.2% of cases. No association was found for any tested variable (age, sex, use of mTOR inhibitors, and multiple tumors) with clinical evolution.

**Conclusions:** Our data indicate a considerable prevalence of arrhythmias and the persistence of identifiable masses throughout follow-up in a brazilian cohort of patients with tuberous sclerosis complex, emphasizing the need for continued cardiological monitoring.

**Clinical Perspective:** *What is new?:*

- Incomplete regression of cardiac rhabdomyomas was the most frequent outcome, occurring in 76.2% of cases.
- A notable prevalence of arrhythmias (21.43%) was observed during follow-up, despite most patients being asymptomatic.

*What are the clinical implications?:*

- The significant occurrence of arrhythmias underscores the necessity for ongoing cardiac monitoring in patients with tuberous sclerosis complex, even when asymptomatic.
- The persistence of identifiable cardiac masses over time highlights the importance of regular imaging studies to assess tumor evolution.

## 1. Introduction

Tuberous Sclerosis Complex (TSC) is a rare genetic disease with an incidence ranging from 1:6.760 to 1:13.520 live births, with no gender predilection^1^. It is an autosomal dominant disorder with variable penetrance, resulting from mutations in the TSC1 (tuberous sclerosis complex-1) or TSC2 (tuberous sclerosis complex-2) genes, which encode the regulatory proteins hamartin and tuberin, respectively^2^.

Neurological and cutaneous manifestations are the most common. Epilepsy is the most frequent symptom, affecting 80–90% of patients, often starting within the first year of life^3^. Cortical tubers, responsible for the disease’s name, occur in up to 90% of cases and serve as potential substrates for epileptic seizures^4^. Hypomelanotic lesions are also present in 90% of TSC patients, usually appearing at birth^5^.

Renal manifestations are the leading cause of morbidity and mortality in adults with TSC^6^. Renal angiomyolipomas are present in 48% of patients^7^ and can lead to hypertension, progressive renal function impairment and an increased risk of hemorrhagic complications^8^.

On the other hand, in patients under 10 years of age, cardiac rhabdomyomas are the most frequent cause of mortality^9^. They are found in 34.3% to 48% of cases^10,11^, and it is estimated that 51% to 86% of rhabdomyomas are associated with TSC^12^.

These tumors tend to develop between the 20th and 30th weeks of gestation, influenced by maternal hormones^9^. They are more common in patients with TSC2 mutations, which may be associated with more severe disease manifestations^11^.

Rhabdomyomas vary significantly in size and are multiple in 90% of cases, typically located in the right and left ventricles with similar distribution^9^. Most regress spontaneously during the first year of life and are observed less frequently after two years of age^11^.

In most cases, they are asymptomatic, but they may cause clinical repercussions soon after birth or within the first year of life^9^. This can occur due to obstruction of the inflow or outflow tracts of the ventricles, leading to significant pressure gradients, myocardial infiltration causing heart failure, or acting as substrates for complex arrhythmias^9^, as shown in Figure 1. In some cases, large tumors can result in significant hemodynamic compromise and even intrauterine death^13^.

**Figure 1.**
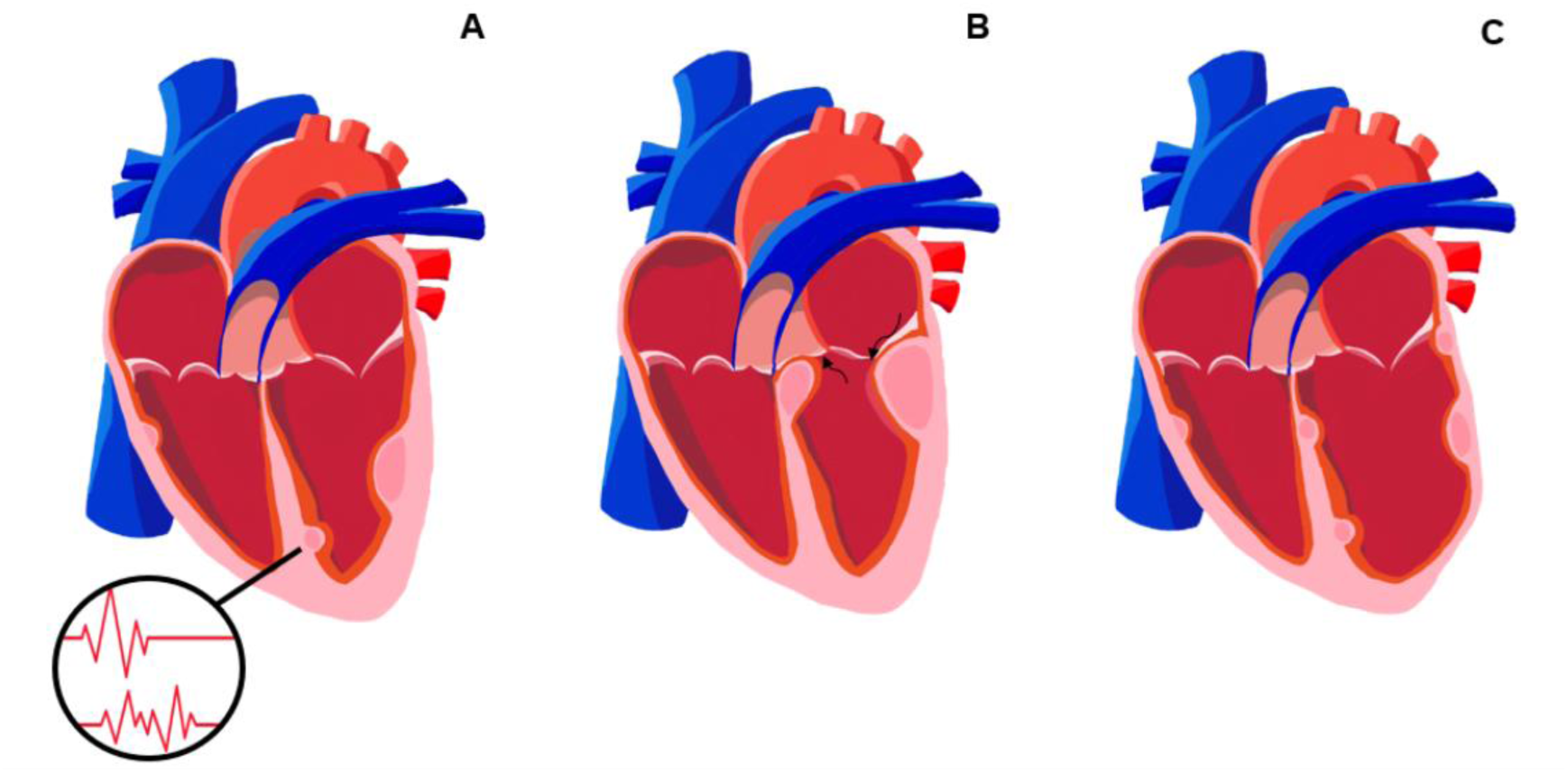
Main manifestations associated with cardiac rhabdomyomas. Representative illustration of the main manifestations associated with cardiac rhabdomyomas. A: Tumor masses as substrates for arrhythmias. B: Obstruction of the inflow and/or outflow tracts of the ventricles with hemodynamic repercussions. C: Heart failure associated with extensive tumor infiltration. Source: Santorio NC (2025).

Transthoracic echocardiography (TTE) is the first-choice imaging method for diagnosis and follow-up due to its wide availability and the possibility of serial assessments, in addition to detecting potential hemodynamic changes. Echocardiography is indicated as early as the fetal stage when compatible abnormalities are observed in morphological ultrasound, in all pediatric patients under three years of age, and in adults with clinical manifestations^14^. In patients diagnosed with cardiac involvement who are asymptomatic, it is recommended to repeat the examination every 1 to 3 years until tumor regression is documented^14^.

Cardiac magnetic resonance imaging (CMR) is considered the gold standard for diagnosing cardiac masses and may be useful in cases of atypical tumor presentation and for surgical planning when indicated^15,16^.

Currently, surgical resection must be considered for tumors causing severe hemodynamic compromise or associated with arrhythmias refractory to standard medications^9^. Mammalian target of rapamycin (mTOR) inhibitors may be considered a therapeutic option in symptomatic patients with hemodynamically significant obstruction associated with the tumor, to avoid surgery or when the surgical procedure is contraindicated^17^.

The diagnostic criteria for TSC were established in 2012^18^ and updated in 2021^14^. These criteria are based on a combination of clinical findings or a positive genetic evaluation. Despite advancements in diagnostic criteria, diagnosis remains challenging, especially in countries like Brazil, due to variability in clinical presentation and limited access to genetic testing, imaging exams, and specialized professionals.

Brazilian studies on tuberous sclerosis remain limited. In 2017, Rosset *et al.* published the first Brazilian study correlating genotype and phenotype in tuberous sclerosis^19^. A total of 53 patients with TSC were evaluated, with pathogenic mutations identified in 89% of cases and rhabdomyomas in 33% of the sample. An overall prevalence 2.5 times higher of TSC2 mutations compared to TSC1 was observed, with no significant differences in the prevalence of cardiac tumors^19^.

Recently, in 2024, Camargo *et al.* published the results of a cohort study of 63 fetuses with cardiac rhabdomyomas from two Brazilian centers, with a median follow-up of 70 months^20^. The overall mortality rate was 17.4%, with 72.7% of deaths occurring during the fetal stage or early neonatal phase^20^.

The present study aims to evaluate clinical outcomes in patients with cardiac tumors, as well as to conduct an evolutionary assessment of cardiac tumors through serial echocardiograms.

## 2. Methods

This is a retrospective, descriptive, single-center study. It included pediatric (0 to 18 years) and adult (over 18 years) patients of both genders with a confirmed diagnosis of TSC based on the updated 2021 criteria^14^, covering the period from January 1997 to January 2024. Patients were selected from the outpatient clinics of Cardiomyopathies, Pulmonology, and Congenital Heart Disease Units of the Heart Institute, Faculty of Medicine, University of São Paulo.

Initially, an electronic search was conducted using the international disease classification in the institution’s electronic medical record system. The data were reviewed independently by two researchers to confirm the diagnosis. Patients with at least two serial TTEs performed at the institution were included, while those with uncertain diagnoses and incomplete records were excluded.

Given the study’s characteristics, individual informed consent was waived while ensuring confidentiality and data protection. The study was approved by the Medical Ethics Committee of the Hospital das Clínicas, Faculty of Medicine, University of São Paulo.

Anthropometric data, gender, ethnicity, and age at diagnosis were collected. Body surface area was calculated using the Haycock formula^21^ for patients under 18 years and the Dubois formula^22^ for those over 18 years.

Regarding systemic involvement, we described the presence of neurological, renal, pulmonary, dermatological, and ophthalmological abnormalities based on the diagnostic criteria defined in the literature^14^ in both groups. Data were also collected on neurological manifestations, malignant neoplasms, the use of mTOR pathway inhibitor medication, and other possible disease-associated manifestations.

For patients with cardiac tumors, clinical data were collected on the presence of dyspnea, chest pain, syncope, and tachycardia/palpitations, in addition to evaluating complementary cardiological exams (TTE, Holter, and CMR).

The first and last available TTE exams were analyzed according to the updated reference values and recommendations of the American Society of Echocardiography^23^. Data collected included cardiac dimensions (anteroposterior diameter of the left atrium, final diastolic diameter of the left ventricle, diastolic measurements of the septum, posterior wall, and aortic sinuses), left ventricular ejection fraction (LVEF) by the Teichholz method, and the presence of pericardial effusion. Right ventricular dimensions were assessed qualitatively, as the measurement window varied significantly between pediatric and adult exams.

For patients who began follow-up between 0 and 18 years of age, the Z-score for echocardiographic measurements was calculated according to Lopez *et al*.^24^ The left atrium diameter was assessed according to the scoring system proposed by Pettersen *et al.*^25^, as it was not covered by the previous reference.

The presence of rhabdomyomas, their quantity (single or multiple tumors), and location (ventricles and/or atria) were characterized, along with hemodynamic repercussions (presence or absence of flow restriction signs in obstructive tumors).

Regarding tumor evolution in serial TTEs, patients were classified as having complete regression, incomplete regression, tumor growth, or recurrence over the follow-up period. Outcomes were described considering the need for surgical treatment, heart transplantation, or all-cause mortality.

Holter-detected abnormalities were described when arrhythmias with a density greater than 1% were observed. Ventricular arrhythmias were classified as simple or complex, with the latter defined by the presence of sustained ventricular tachycardia, non-sustained ventricular tachycardia, or ventricular fibrillation. In CMR analysis, the presence of myocardial fibrosis was evaluated using the late enhancement technique. When available, genetic study data on identified mutations were also collected.

Using the taxpayer identification registry recorded in medical records, a search was conducted in the Brazilian Federal Revenue system to verify patient mortality with and without cardiac involvement, allowing for general mortality data estimation.

### Statistical Analysis

Normality was tested using the Kolmogorov-Smirnov test for all quantitative variables. For normally distributed variables, descriptive statistics were presented as means and standard deviations (mean ± SD), while non-normally distributed variables were reported as medians and interquartile ranges (IQR, 25%-75%). Qualitative variables were expressed as absolute (n) and relative (%) frequencies.

To test associations between unpaired qualitative/categorical variables, Fisher’s exact test or the Chi-square test was used. For paired categorical variable comparisons, McNemar’s asymptotic test was applied for two-category responses, and the Stuart-Maxwell marginal homogeneity test was used for variables with more than two categories.

Comparisons of unpaired quantitative data were performed using the Student’s t-test for normally distributed variables or the Wilcoxon-Mann-Whitney test for non-normally distributed variables. For the variable “age,” which did not follow a normal distribution, the comparison between the three outcome groups was conducted using the Kruskal-Wallis test. Paired quantitative data were compared using a paired t-test or Wilcoxon paired test.

An analysis was performed to evaluate the association between clinical progression and certain individual variables. Additionally, a multivariate analysis using ordinal proportional odds logistic regression^26^ was conducted to identify potential factors associated with unfavorable rhabdomyoma progression.

A significance level of 5% was adopted for all statistical tests, which were considered two-tailed. All statistical calculations were performed using R software version 4.2.2 (R Core Team, 2022) by an independent statistical service.

## 3. Results

Initially, 89 medical records of patients diagnosed with TSC were included. Of these, 20 records were excluded due to incomplete data or uncertain diagnosis.

Among the 69 evaluated patients, 42 (60.9%) had cardiac tumors, including 41 rhabdomyomas (Figure 2) and one pericardial lipoma (Figure 3). The demographic variables of both groups are presented in Table 1.

**Figure 2.**
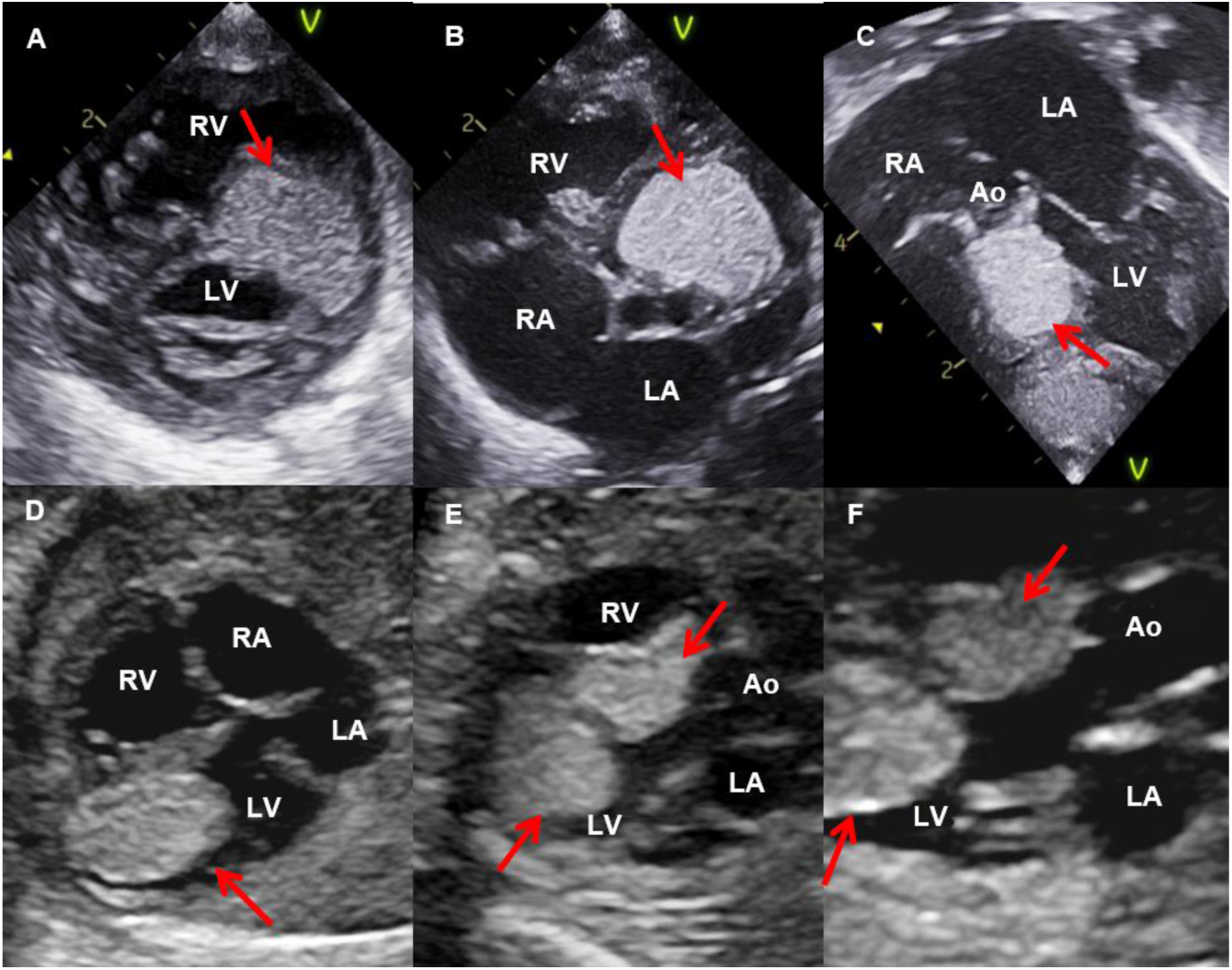
Cardiac rhabdomyomas observed by transthoracic and fetal echocardiography. In A, B, and C, transthoracic echocardiography images show an echogenic mass (arrows) adhered to the interventricular septum, invading and causing obstruction of the left ventricular outflow tract. These findings are demonstrated in the parasternal short-axis view of the ventricles (A), short-axis view of the aortic valve (B), and apical five-chamber view (C). In D, E, and F, fetal echocardiography images reveal echogenic masses (arrows) attached to the interventricular septum and the apical region of the left ventricle. These are shown in the four-chamber view (D) and five-chamber views (E and F). No obstruction of the left ventricular inflow and outflow tracts was observed, with preserved aortic valve opening (F). RA: right atrium; LA: left atrium; RV: right ventricle; LV: left ventricle; Ao: aorta. Exams performed at the Pediatric Echocardiography Department of the Heart Institute.

**Figure 3.**
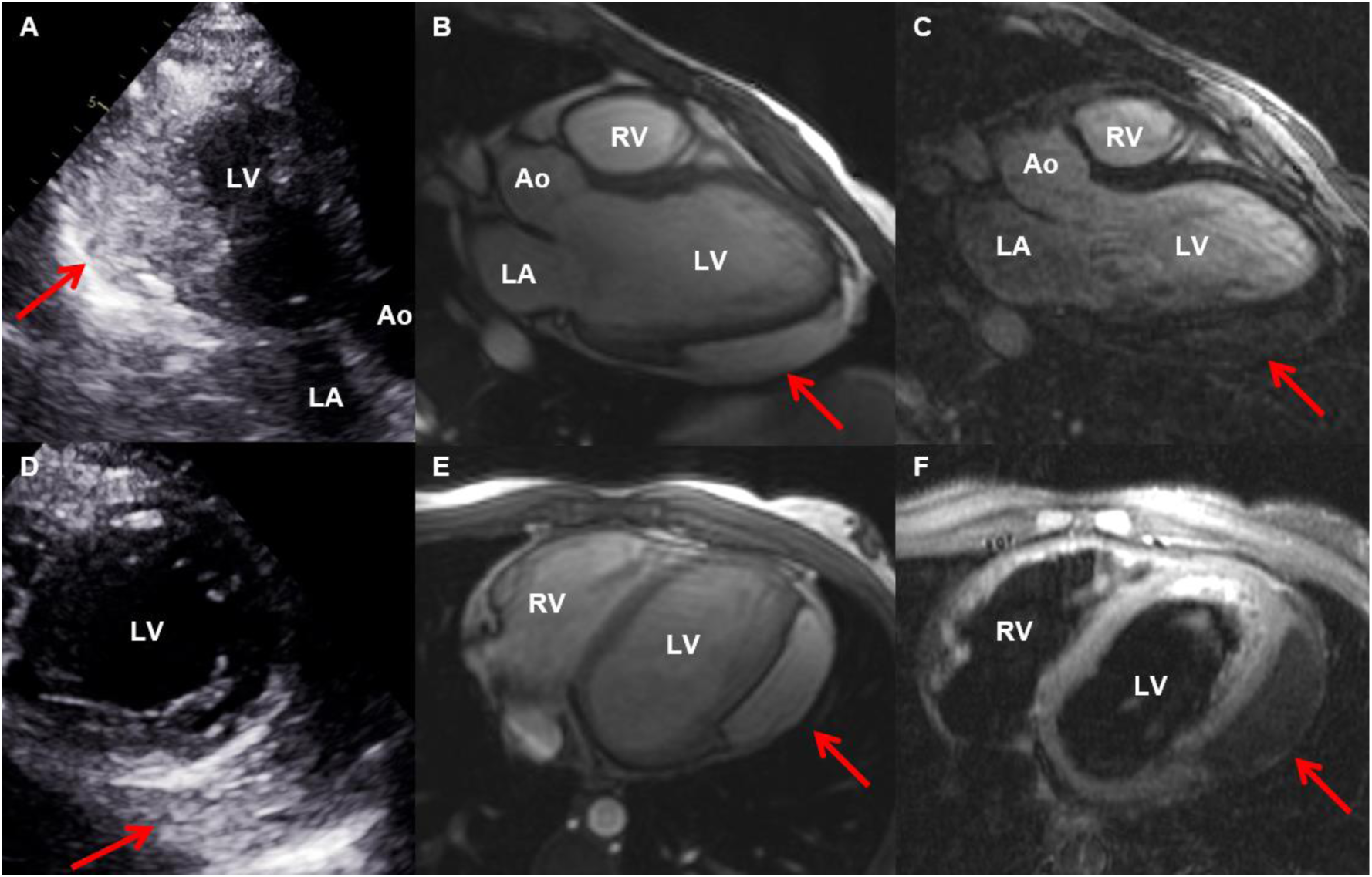
Pericardial lipoma observed by transthoracic echocardiography and cardiac magnetic resonance imaging. Transthoracic echocardiography (A and D) and cardiac magnetic resonance imaging (B, C, E, F) showing a lesion (arrows) in the pericardial region, in close contact with the lateral wall of the left ventricle. In transthoracic echocardiography, a hyperechogenic mass is visualized in the apical three-chamber view (A) and the parasternal short-axis view focused on the left ventricle (D). Cardiac magnetic resonance imaging reveals a lesion with an adipose component in the three-chamber views (B and C) and the longitudinal axis of the ventricles (E and F). No late contrast enhancement is observed (C). In F, a hypointense signal is noted in the T2-weighted fat-saturation sequence, suggesting a lesion with a fatty component. LA: left atrium; RV: right ventricle; LV: left ventricle; Ao: aorta. Exams performed at the Heart Institute.

**Table 1.** Demographic variables of the overall sample and patients with cardiac tumors.

|  | Overall sample (n = 69) | Cardiac tumor sample (n = 42) | No cardiac tumor sample (n = 27) | p (< 0.05) |
| --- | --- | --- | --- | --- |
| Female sex (n, %) | 43 (62.3%) | 21 (50%) | 22 (81.5%) | 0.11<sup>(1)</sup> |
| Age at diagnosis (median [Q1; Q3]) | 4.0 [0.0; 22.0] | 0.5 [0.0; 6.2] | 22.0 [13.5; 33.0] | < **0.001**<sup>(2)</sup> |
| Ethnicity |  |  |  | 0.929<sup>(3)</sup> |
| White (n, %) | 63 (91.3%) | 39 (92.9%) | 24 (88.9%) |  |
| Mixed (n, %) | 3 (4.3%) | 2 (4.8%) | 1 (3.7%) |  |
| Asian (n,%) | 2 (2.9%) | 1 (2.4%) | 1 (3.7%) |  |
| Black (n,%) | 0 (0%) | 0 (0%) | 0 (0%) |  |
| Not reported (n,%) | 1 (1.5%) | 0 (0%) | 1 (3.7%) |  |

(1) Fisher's exact test, (2) Wilcoxon-Mann-Whitney test, (3) Chi-square test

The presence of subependymal nodules was the most common major criterion in patients with cardiac tumors, observed in 85.7% of cases (Table 2). In patients without cardiac tumors, renal angiomyolipomas were more frequent (92.6%). Lymphangioleiomyomatosis (LAM), periungual fibromas, renal and hepatic angiomyolipomas were significantly more prevalent in the group without cardiac tumors.

**Table 2.** Major diagnostic criteria in patients with and without cardiac tumors.

| Major criteria | Total sample<br>(n = 69) | With cardiac tumors<br>(n = 42) | Without cardiac tumors<br>(n = 27) | p (< 0.05) |
| --- | --- | --- | --- | --- |
| Hypomelanotic macules (n,%) | 33 (47.8%) | 20 (47.6%) | 13 (48.2%) | > 0.999 |
| Angiofibroma or fibrous cephalic plaque (n,%) | 41 (59.4%) | 21 (50%) | 20 (74.1%) | 0.078 |
| Periungual fibromas (n, %) | 22 (31.9%) | 7 (16.7%) | 15 (55.6%) | **0.001** |
| Shagreen patches (n, %) | 12 (17.4%) | 6 (14.3%) | 6 (22.2%) | 0.518 |
| Retinal hamartomas (n, %) | 7 (10.1%) | 4 (9.5%) | 3 (11.1%) | > 0.999 |
| Cortical tubers (n, %) | 49 (71%) | 31 (73.8%) | 31 (73.8%) | 0.592 |
| Radial migration lines (n, %) | 24 (34.8%) | 15 (35.7%) | 9 (33.3%) | > 0.999 |
| Subependymal nodules (n,%) | 56 (81.2%) | 36 (85.7%) | 20 (74.1%) | 0.344 |
| SEGA (n,%) | 12 (17.4%) | 8 (19.1%) | 4 (14.8%) | 0.753 |
| Cardiac rhabdomyoma (n,%) | 41 (59.4%) | 41 (97.6%) | 0 (0%) | **< 0.001** |
| LAM (n,%) | 19 (27.5%) | 3 (7.1%) | 16 (59.3%) | **< 0.001** |
| Angiomyolipomas (n,%) |  |  |  |  |
| Renal | 47 (68.1%) | 22 (52.4%) | 25 (92.6%) | 0.002 |
| Hepatic | 11 (15.9%) | 4 (9.5%) | 7 (25.9%) | 0.011 |
| Soft tissue | 1 (1.5%) | 1 (2.4%) | 0 (0%) | - |
| Pancreatic | 1 (1.5%) | 0 (0%) | 1 (3.7%) | - |

SEGA: Subependymal Giant Cell Astrocytoma; LAM: Lymphangioleiomyomatosis. All comparisons were performed using Fisher's Exact Test.

Regarding minor criteria (Table 3), sclerotic bone lesions were the most frequent findings in both groups, present in 33.3% of patients with cardiac involvement and 63% of those without. The presence of intraoral fibromas and sclerotic bone lesions was significantly more frequent in the group without cardiac tumors.

**Table 3.**
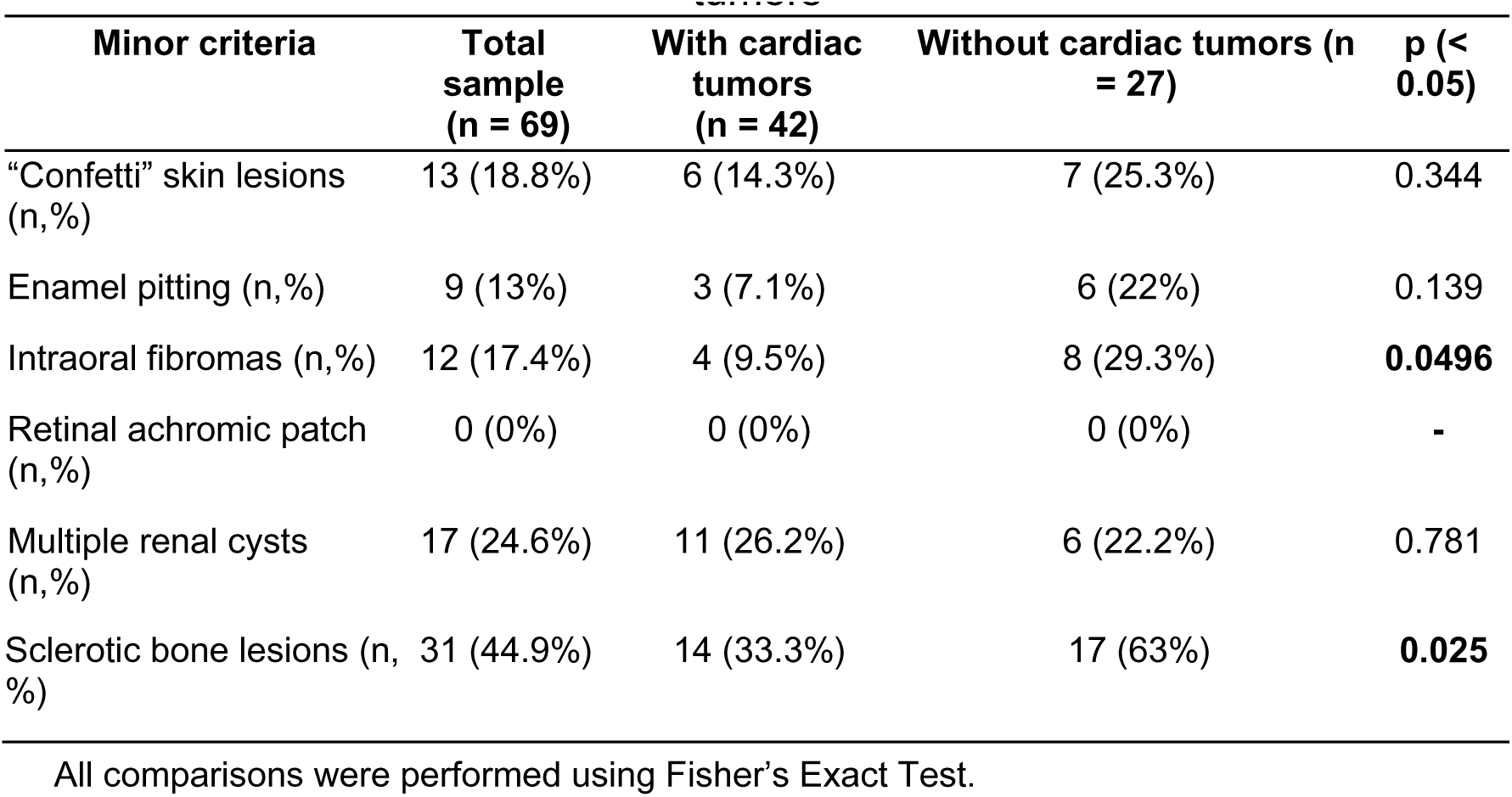
Minor diagnostic criteria present in patients with and without cardiac tumors.

| Minor criteria | Total sample (n = 69) | With cardiac tumors (n = 42) | Without cardiac tumors (n = 27) | p (< 0.05) |
| --- | --- | --- | --- | --- |
| “Confetti” skin lesions (n,%) | 13 (18.8%) | 6 (14.3%) | 7 (25.3%) | 0.344 |
| Enamel pitting (n,%) | 9 (13%) | 3 (7.1%) | 6 (22%) | 0.139 |
| Intraoral fibromas (n,%) | 12 (17.4%) | 4 (9.5%) | 8 (29.3%) | **0.0496** |
| Retinal achromic patch (n,%) | 0 (0%) | 0 (0%) | 0 (0%) | - |
| Multiple renal cysts (n,%) | 17 (24.6%) | 11 (26.2%) | 6 (22.2%) | 0.781 |
| Sclerotic bone lesions (n, %) | 31 (44.9%) | 14 (33.3%) | 17 (63%) | **0.025** |

All comparisons were performed using Fisher’s Exact Test.

On the other hand, seizures were more commonly reported in patients with cardiac tumors (90.5% versus 51.9%, p < 0.001) and were present in 75.4% of the overall sample. Delayed neuropsychomotor development (35.7% vs. 22.2%, p = 0.23) and autism spectrum disorder (11.9% vs. 14.8%, p = 0.73) did not show significant differences between the groups.

In the gender-based comparative analysis, rhabdomyomas (76.9% vs. 48.8%, p = 0.02) were significantly more prevalent in males. Seizures (92.3% vs. 65.1%, p = 0.011) and delayed neuropsychomotor development (46.2% vs. 20.9%, p = 0.027) were also more frequent in males.

Genetic analysis was obtained for only three patients (4.3% of the sample), identifying two mutations in the TSC2 gene and one negative genetic test.

All-cause mortality data were obtained and confirmed for 89.9% of the sample. In the group with cardiac tumors, the mortality rate was 7.1%, while in the group without cardiac tumors, it was 8.3% (p > 0.999).

The occurrence of malignant neoplasms also showed no significant difference between the groups, although it was numerically higher in the group without cardiac tumors (11.1% vs. 4.8%, p = 0.37). The use of mTOR inhibitors was more frequent in the group without cardiac tumors (44.4% vs. 14.3%, p = 0.005) and notably higher in females (37.2% vs. 7.69%, p < 0.01).

Regarding clinical manifestations in patients with cardiac tumors, the vast majority of patients were asymptomatic in both evaluations (73.8% and 85.7%, respectively). Dyspnea was the most frequently reported symptom in both assessments (Table 4).

**Table 4.**
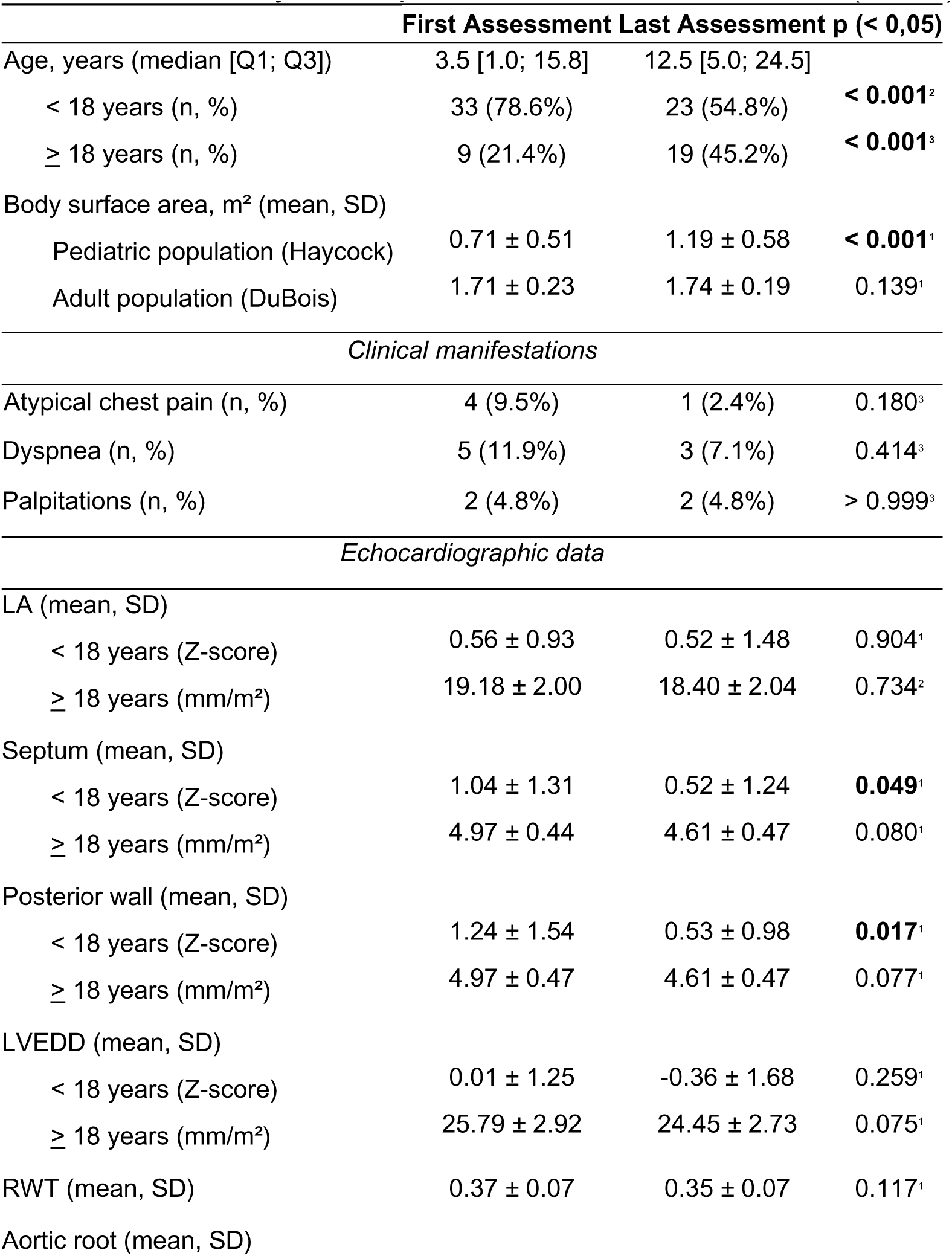
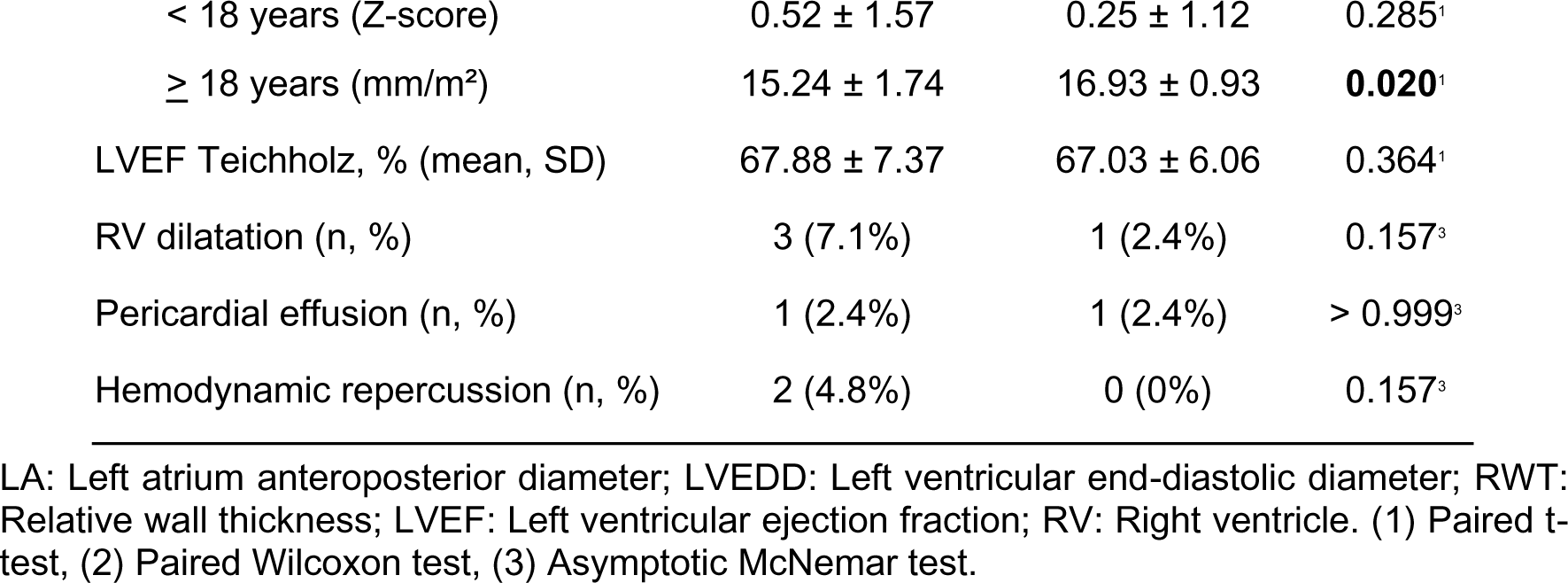
Evolutionary data of patients with cardiac involvement (n = 42).

|  | First Assessment | Last Assessment | p (< 0,05) |
| --- | --- | --- | --- |
| Age, years (median [Q1; Q3]) | 3.5 [1.0; 15.8] | 12.5 [5.0; 24.5] |  |
| < 18 years (n, %) | 33 (78.6%) | 23 (54.8%) | **< 0.001<sup>2</sup>** |
| ≥ 18 years (n, %) | 9 (21.4%) | 19 (45.2%) | **< 0.001<sup>3</sup>** |
| Body surface area, m<sup>2</sup> (mean, SD) |  |  |  |
| Pediatric population (Haycock) | 0.71 ± 0.51 | 1.19 ± 0.58 | **< 0.001<sup>1</sup>** |
| Adult population (DuBois) | 1.71 ± 0.23 | 1.74 ± 0.19 | 0.139<sup>1</sup> |
| *Clinical manifestations* |  |  |  |
| Atypical chest pain (n, %) | 4 (9.5%) | 1 (2.4%) | 0.180<sup>3</sup> |
| Dyspnea (n, %) | 5 (11.9%) | 3 (7.1%) | 0.414<sup>3</sup> |
| Palpitations (n, %) | 2 (4.8%) | 2 (4.8%) | > 0.999<sup>3</sup> |
| *Echocardiographic data* |  |  |  |
| LA (mean, SD) |  |  |  |
| < 18 years (Z-score) | 0.56 ± 0.93 | 0.52 ± 1.48 | 0.904<sup>1</sup> |
| ≥ 18 years (mm/m<sup>2</sup>) | 19.18 ± 2.00 | 18.40 ± 2.04 | 0.734<sup>2</sup> |
| Septum (mean, SD) |  |  |  |
| < 18 years (Z-score) | 1.04 ± 1.31 | 0.52 ± 1.24 | **0.049<sup>1</sup>** |
| ≥ 18 years (mm/m<sup>2</sup>) | 4.97 ± 0.44 | 4.61 ± 0.47 | 0.080<sup>1</sup> |
| Posterior wall (mean, SD) |  |  |  |
| < 18 years (Z-score) | 1.24 ± 1.54 | 0.53 ± 0.98 | **0.017<sup>1</sup>** |
| ≥ 18 years (mm/m<sup>2</sup>) | 4.97 ± 0.47 | 4.61 ± 0.47 | 0.077<sup>1</sup> |
| LVEDD (mean, SD) |  |  |  |
| < 18 years (Z-score) | 0.01 ± 1.25 | -0.36 ± 1.68 | 0.259<sup>1</sup> |
| ≥ 18 years (mm/m<sup>2</sup>) | 25.79 ± 2.92 | 24.45 ± 2.73 | 0.075<sup>1</sup> |
| RWT (mean, SD) | 0.37 ± 0.07 | 0.35 ± 0.07 | 0.117<sup>1</sup> |
| Aortic root (mean, SD) |  |  |  |
| < 18 years (Z-score) | 0.52 ± 1.57 | 0.25 ± 1.12 | 0.285<sup>1</sup> |
| ≥ 18 years (mm/m<sup>2</sup>) | 15.24 ± 1.74 | 16.93 ± 0.93 | **0.020<sup>1</sup>** |
| LVEF Teichholz, % (mean, SD) | 67.88 ± 7.37 | 67.03 ± 6.06 | 0.364<sup>1</sup> |
| RV dilatation (n, %) | 3 (7.1%) | 1 (2.4%) | 0.157<sup>3</sup> |
| Pericardial effusion (n, %) | 1 (2.4%) | 1 (2.4%) | > 0.999<sup>3</sup> |
| Hemodynamic repercussion (n, %) | 2 (4.8%) | 0 (0%) | 0.157<sup>3</sup> |

LA: Left atrium anteroposterior diameter; LVEDD: Left ventricular end-diastolic diameter; RWT: Relative wall thickness; LVEF: Left ventricular ejection fraction; RV: Right ventricle. (1) Paired t-test, (2) Paired Wilcoxon test, (3) Asymptotic McNemar test.

In the group of patients with cardiac tumors, episodes of supraventricular and ventricular tachycardia were recorded in 16.7% and 7.2% of the sample, respectively. Wolff-Parkinson-White syndrome was described in 9.5% of cases. Overall, arrhythmias occurred in 21.4% of the sample at some point during follow-up.

Regarding medication use, 23.8% of patients used beta-blockers, 14.3% used renin-angiotensin-aldosterone system inhibitors, 11.9% used diuretics, and 7.1% used amiodarone.

The median follow-up period between the first and last TTE performed was 6 years [2.2; 9.8]. Throughout the follow-up period, there was no clinically significant change in the echocardiographic data analyzed (Table 4). Only one patient (2.4%) had LVEF < 50%.

The isolated involvement of the left ventricle and biventricular involvement were the most common locations for rhabdomyomas (43.9% each). Biventricular involvement was the most frequent presentation in patients with multiple rhabdomyomas (58.1%), as shown in Table S1.

A 24-hour Holter monitor was performed in 20 patients, revealing the following rhythms: sinus rhythm (60%), sinus rhythm with atrial ectopy (35%), and sinus rhythm with sinus arrhythmia (5%). Simple and complex arrhythmias were present in 25% and 5% of the sample, respectively.

CMR was available for 15 patients with cardiac tumors, with 20% of them presenting late gadolinium enhancement (LGE). In only 13.3% of these cases, tumors were identified exclusively by CMR and were not detected in the initial TTE.

The most frequent presentation was incomplete mass regression in 76.2% of cases, with complete regression occurring in 16.7% (Figure S1). Only one patient (2.4%) showed tumor growth, and another had stable tumor size. Surgical treatment was required in one case, which later resulted in death during a separate hospitalization due to difficult-to-control ventricular arrhythmias (2.4%). No patient underwent heart transplantation or received an implantable cardioverter-defibrillator, while three patients (7.14%) underwent catheter ablation.

The patient with pericardial lipoma maintained tumor size stability between evaluations but required ablation for epicardial ventricular tachycardia during follow-up, remaining asymptomatic afterward.

An association between clinical progression and selected variables was analyzed (Table 5). Among the analyzed variables, patients with tumor growth, need for surgical treatment, and/or death had a higher prevalence of complex arrhythmias on Holter monitoring (p = 0.051). However, it was not possible to correlate complex arrhythmias on Holter with LGE on CMR due to the small number of available exams in the sample.

**Table 5.** Association of outcomes with selected sociodemographic and clinical variables among patients with cardiac tumors.

|  | **Complete**<br>**regression**<br>**(n = 7)** | **Incomplete**<br>**regression**<br>**(n = 32)** | **Mass maintenance,**<br>**increase, or surgical**<br>**treatment**<br>**(n = 3)** | **p (<**<br>**0.05)** |
| --- | --- | --- | --- | --- |
| Age, years (median [Q1; Q3]) | 1.0 [0.5; 1.5] | 0.0 [0.0; 9.2] | 0.0 [0.0; 7.5] | 0.952 |
| Female sex (n, %) | 4 (57.1%) | 16 (50.0%) | 1 (33.3%) | 0.788 |
| Multiple tumors | 5 (71.4%) | 24 (75.0%) | 2 (66.7%) | 0.940 |
| LVEF < 50% at presentation | 0 (0.0%) | 1 (3.1%) | 0 (0.0%) | - |
| Late gadolinium enhancement on CMR | 1 (50%) | 1 (9.1%) | 1 (50%) | 0.216 |
| Complex arrhythmias on Holter | 0 (0%) | 0 (0%) | 1 (33.3%) | 0.051 |
| Use of mTOR inhibitor | 1 (14.3%) | 4 (12.5%) | 1 (33.3%) | 0.615 |

LVEF: Left ventricular ejection fraction; CMR: Cardiac magnetic resonance; mTOR: Mammalian target of rapamycin.

The ordinal logistic regression analysis of proportional odds showed no significant association between any tested variables and clinical progression (Table S2).

## 4. Discussion

This study provides a detailed and comprehensive analysis of the clinical manifestations, systemic involvement, and progression of patients with rhabdomyomas associated with TSC in a brazilian cohort.

Classically, as an autosomal dominant disease, there is no significant difference in the prevalence of TSC between genders. In the overall sample, a slight predominance of females (62.3%) was observed, mainly due to a significantly higher prevalence in the group without cardiac involvement (81.5%). This finding is possibly attributed to the high prevalence of patients with LAM (59.3%) in the group without cardiac tumors, considering that our center also specializes in pulmonology.

Another interesting aspect was the lower prevalence of rhabdomyomas in females, which is not described in the literature^10,11^. A possibly associated factor with this finding is the higher proportion of mTOR inhibitor use among females in our sample (37.2% versus 7.7%). In 2021, Chen *et al*. described increased rates of cardiac rhabdomyoma resolution with mTOR inhibitors, with greater efficacy in women and younger patients^27^. It is possible, therefore, that female patients experienced a faster reduction in tumor mass, making them undetectable by transthoracic echocardiograms at the beginning of follow-up in our service.

On the other hand, female gender and mTOR inhibitor use were not associated with higher rates of complete rhabdomyoma regression in our study. This finding may be understood in the perspective that, in patients with a previous diagnosis, examiners may be more inclined to actively search for and consider small lesions that may go unnoticed in patients without a prior diagnosis. Additionally, such patients are more frequently subjected to CMR exams, which have higher sensitivity for detecting small lesions.

Another relevant finding was the significantly younger age at diagnosis in the group with cardiac involvement. This finding, already reported in other centers^1^, is likely due to the earlier diagnosis of rhabdomyomas, often made during the prenatal period through fetal echocardiography. Furthermore, rhabdomyomas have been described as the most frequent manifestation in children up to 36 months old, present in 59% of cases^28,29^.

The prevalence of rhabdomyomas in our institution was 59.4%, higher than the literature-reported range^10,11^. This difference is likely associated with the fact that we are a national reference center in pediatric and adult cardiology, receiving cases with suspected cardiac involvement. Therefore, the population of patients without cardiac tumors was probably underrepresented.

The presentation of a pericardial lipoma associated with TSC is quite rare, with no prior descriptions in the literature. It was recorded in a male patient who presented with ventricular tachycardia and maintained tumor size in serial evaluations.

Regarding other manifestations, the overall occurrence was similar to that reported in other centers, except for a lower occurrence of cutaneous, oral, retinal, and sclerotic bone lesions^30^. These data may be underestimated, as they were based on medical records filled out by cardiologists and pulmonologists, who may not have actively investigated or inquired about these manifestations during follow-up.

When comparing groups with and without cardiac involvement, seizures were significantly more prevalent in patients with cardiac tumors (90.5% versus 51.9%, p = 0.0003). In 2016, Jeong and Wong showed that rhabdomyomas were the only systemic manifestation that remained significantly associated with epilepsy in multivariate analysis^31^, aligning with our findings. However, there were no differences in identifiable anatomical lesions in neuroimaging exams. This finding suggests that the pathophysiology of seizures is not yet fully elucidated, and further studies are necessary to clarify the mechanisms behind this possible correlation.

Recently, in 2022, Samuel YL *et al.* reported a higher occurrence of cardiac rhabdomyomas, renal angiomyolipomas, and facial angiofibromas in Chinese patients with TSC2 mutations, suggesting a possible association between cardiac tumors and more severe systemic involvement^32^. This association was not found in our study, as systemic manifestations such as renal angiomyolipomas, LAM, sclerotic bone lesions, and intraoral and periungual fibromas were more frequent in the group without cardiac involvement. Genetic analysis would certainly provide relevant insights into the genotype-phenotype correlation in our population. However, due to limited availability in the public healthcare system, only 4.3% of the sample underwent genetic testing, restricting data interpretation.

Regarding all-cause mortality, the group without cardiac tumors showed a trend toward higher all-cause mortality (8.3% versus 7.1%), though not statistically significant. A 2017 study showed that renal disease is the leading cause of mortality in patients with TSC^33^. Unfortunately, specific mortality cause data were not obtained in our study, but the group without cardiac involvement had a notably higher prevalence of renal angiomyolipomas, which could be related to these findings.

In our sample, cardiac tumors were predominantly multiple in 75.6% of cases, a proportion slightly lower than the 90% reported in other studies^9^. This may be associated with the predominant use of transthoracic echocardiography in our study.

Most patients with cardiac rhabdomyomas remained asymptomatic during follow-up (73.8% at the first and 85.71% at the last evaluation). In 2006, Jóźwiak *et al*. evaluated 74 patients with TSC-associated rhabdomyomas, showing that 61% of tumors had no clinical manifestations, with only one death directly related to cardiac involvement (1.4%) and a 5.4% incidence of heart failure^11^. In 1996, Bosi *et al*. reported only one case of heart failure (3.3%) in a series of 30 TSC-associated rhabdomyomas, with no deaths directly related to cardiac involvement and no tumor-related surgical indications^34^. Our data also indicated a rare occurrence of these outcomes, with only one heart failure case, one death, and one surgical intervention in the same patient (2.4%).

Jóźwiak *et al.* reported a 23% arrhythmia rate^11^, while Bosi et al. found a 26.7% rate^34^. Our study had similar values (21.4% arrhythmias, with 16.7% supraventricular tachycardias and 7.2% ventricular tachycardias). Nir *et al.* described Wolff-Parkinson-White syndrome in 9% of patients with rhabdomyomas, also consistent with our findings (9.5%)^35^.

Until now, no longitudinal studies have evaluated the progression of TSC-associated rhabdomyomas in the Brazilian population. Our study showed a complete regression rate lower than that of more recent studies^10,36^, at only 16.7%, with incomplete regression being the most common clinical outcome (76.2%). This difference from older studies is likely due to advancements in imaging techniques, which allow for greater sensitivity in detecting smaller tumors. Furthermore, adult patients were also included, whereas complete regression is classically described in early childhood.

Finally, the median follow-up period was six years, primarily determined by the availability of electronic medical records. While this timeframe likely suffices for characterizing early tumor evolution, longer follow-up studies could be useful in assessing possible tumor growth or new tumor development during puberty.

Our study presents limitations. The availability of data was conditioned by electronic records made by different professionals over 27 years. During this period, there was an improvement in imaging techniques and a progressive migration to more computerized systems. Undoubtedly, the application of standardized questionnaires would provide more accurate and uniform information on systemic and cardiological manifestations in patients with TSC.

The evolutionary assessment of rhabdomyomas was primarily performed through comparisons between transthoracic echocardiograms conducted during follow-up. However, the examinations were performed by different operators from distinct teams (adult and pediatric echocardiography). Additionally, these patients were evaluated using the Z-score calculation for linear measurements throughout the follow-up, even if they were over 18 years old at the last examination. Although this methodology is not validated, it was used to facilitate the evolutionary comparison of variables, which may have impacted the results.

Furthermore, although CMR is considered the gold standard for diagnosing rhabdomyomas, this study included a pediatric population. This makes it unfeasible to perform the exam in all patients diagnosed with cardiac tumors due to the need for sedation. Therefore, inferences related to the presence of late enhancement in tumor prognosis were limited by the small number of examinations performed.

Another important limitation was the low availability of genetic testing, which prevents the establishment of a genotype-phenotype correlation in the studied population.

Additionally, this is a single-center retrospective study, which may not represent the Brazilian population in general. Given that TSC is a disease with a pathophysiology that is not yet fully understood, studies in specific populations could reveal particularities associated with interactions between genetics and environmental, nutritional, racial, and epidemiological aspects. However, due to the rarity of this disease, limited resources, and challenges in making a definitive diagnosis, there are multiple obstacles to conducting multicenter studies on this topic in Brazil.

In this study, low mortality rates of only 2.4% were found. The Brazilian fetal tumor cohort published in 2024 by Camargo *et al*. reported a higher mortality rate of 17.4%^20^. However, most deaths (72.7%) occurred in the fetal or early neonatal period. Since our center does not have its own maternity service and does not provide prenatal follow-up, most patients with cardiac tumors began follow-up at an outpatient level after the initial peak of reported deaths, which justifies the discrepancy in rates found.

Although mortality significantly decreases after the neonatal period, most patients retained identifiable tumors in sequential examinations, representing potential arrhythmogenic substrates. In agreement with this, a significant rate of cardiac arrhythmias was found, reinforcing the importance of specialized longitudinal follow-up in cardiology.

Concerning treatment, the use of mTOR inhibitors did not show an association with improved clinical outcomes. However, this finding should be interpreted with caution, considering the low occurrence of heart failure, death, and the need for surgical intervention in the sample. Randomized studies may clarify the potential benefits of these medications for cardiac rhabdomyomas, which are currently reserved for symptomatic cases or large-volume masses.

## 5. Conclusions

In this cohort, incomplete remission was the most frequent outcome of cardiac tumors evaluated through serial echocardiograms. Cardiac tumors were associated with a considerable occurrence of arrhythmias during follow-up, exceeding 20%, despite low mortality rates and limited surgical treatment.

Regarding systemic manifestations, cardiac rhabdomyomas were associated with a higher occurrence of seizures. On the other hand, renal angiomyolipomas, LAM, sclerotic bone lesions, intraoral and periungual fibromas were more frequent in the group without cardiac involvement.

No factors were found to be associated with the progression of cardiac tumors among the variables analyzed in the sample (use of mTOR inhibitors, presence of late gadolinium enhancement on CMR, LVEF < 50% at presentation, female sex, age, and multiple tumors). Further studies with larger samples and longer follow-up periods are needed to clarify the impact of these variables.

## Data Availability

All data generated or analyzed during this study are included in this published article [and its supplementary information files]. Additional datasets used and/or analyzed during the current study are available from the corresponding author on reasonable request.

## Acknowledgments

We extend our gratitude to the teams of Congenital Heart Disease, Pulmonology and Cardiomyopathies at the Heart Institute for their unwavering dedication and efforts in providing high-quality treatment and continuous follow-up for patients with tuberous sclerosis.

## Supplemental Material

Tables S1–S2

Figure S1

## References

1. Ebrahimi-Fakhari D, Meyer S, Vogt T, Pföhler C, Müller CSL. Dermatological manifestations of tuberous sclerosis complex (TSC). JDDG: Journal der Deutschen Dermatologischen Gesellschaft. 2017;15:695–700.

2. Crino PB, Nathanson KL, Henske EP. The tuberous sclerosis complex. New England Journal of Medicine. 2006;355:1345–1356.

3. Henske EP, Jóźwiak S, Kingswood JC, Sampson JR, Thiele EA. Tuberous sclerosis complex. Nature reviews Disease primers. 2016;2:1–18.

4. Valente KD, Sampaio LB, Vincentiis S, Pinto ALR, Montenegro MA. Tuberous Sclerosis Complex: An updated in the treatment of epilepsy for early careers. Epilepsy & Behavior. 2025;167:110396.

5. Cascarino M, Leclerc-Mercier S. Histological patterns of skin lesions in tuberous sclerosis complex: a panorama. Dermatopathology. 2021;8:236–252.

6. Trnka P, Kennedy SE. Renal tumors in tuberous sclerosis complex. Pediatric Nephrology. 2021;36:1427–1438.

7. Kingswood JC, Belousova E, Benedik MP, Carter T, Cottin V, Curatolo P, Dahlin M, D’Amato L, Beaure d’Augères G, de Vries PJ. Renal manifestations of tuberous sclerosis complex: key findings from the final analysis of the TOSCA study focussing mainly on renal angiomyolipomas. Frontiers in neurology. 2020;11:972.

8. Caban C, Khan N, Hasbani DM, Crino PB. Genetics of tuberous sclerosis complex: implications for clinical practice. The application of clinical genetics. 2016:1–8.

9. Hinton RB, Prakash A, Romp RL, Krueger DA, Knilans TK. Cardiovascular manifestations of tuberous sclerosis complex and summary of the revised diagnostic criteria and surveillance and management recommendations from the International Tuberous Sclerosis Consensus Group. Journal of the American Heart Association. 2014;3:e001493.

10. Kingswood JC, d’Augères GB, Belousova E, Ferreira JC, Carter T, Castellana R, Cottin V, Curatolo P, Dahlin M, De Vries PJ. TuberOus SClerosis registry to increase disease Awareness (TOSCA)–baseline data on 2093 patients. Orphanet journal of rare diseases. 2017;12:1-13.

11. Jóźwiak S, Kotulska K, Kasprzyk-Obara J, Domańska-Pakieła D, Tomyn-Drabik M, Roberts P, Kwiatkowski D. Clinical and genotype studies of cardiac tumors in 154 patients with tuberous sclerosis complex. Pediatrics. 2006;118:e1146–e1151.

12. Harding CO, Pagon RA. Incidence of tuberous sclerosis in patients with cardiac rhabdomyoma. American journal of medical genetics. 1990;37:443–446.

13. Groves A, Fagg N, Cook AC, Allan LD. Cardiac tumours in intrauterine life. Archives of disease in childhood. 1992;67:1189–1192.

14. Northrup H, Aronow ME, Bebin EM, Bissler J, Darling TN, de Vries PJ, Frost MD, Fuchs Z, Gosnell ES, Gupta N. Updated international tuberous sclerosis complex diagnostic criteria and surveillance and management recommendations. Pediatric Neurology. 2021;123:50–66.

15. Freedberg RS, Kronzon I, Rumancik WM, Liebeskind D. The contribution of magnetic resonance imaging to the evaluation of intracardiac tumors diagnosed by echocardiography. Circulation. 1988;77:96–103.

16. Berkenblit R, Spindola-Franco H, Frater RW, Fish BB, Glickstein JS. MRI in the evaluation and management of a newborn infant with cardiac rhabdomyoma. The Annals of thoracic surgery. 1997;63:1475–1477.

17. Dahdah N. Everolimus for the treatment of tuberous sclerosis complex– related cardiac rhabdomyomas in pediatric patients. The Journal of pediatrics. 2017;190:21–26. e27.

18. Northrup H, Krueger DA, Roberds S, Smith K, Sampson J, Korf B, Kwiatkowski DJ, Mowat D, Nellist M, Povey S. Tuberous sclerosis complex diagnostic criteria update: recommendations of the 2012 International Tuberous Sclerosis Complex Consensus Conference. Pediatric neurology. 2013;49:243–254.

19. Rosset C, Vairo F, Bandeira IC, Correia RL, De Goes FV, Da Silva RTB, Bueno LSM, de Miranda Gomes MCS, Galvão HdCR, Neri JI. Molecular analysis of TSC1 and TSC2 genes and phenotypic correlations in Brazilian families with tuberous sclerosis. PLoS One. 2017;12:e0185713.

20. Camargo FM, Brizot MdL, Francisco RPV, Carvalho WBd, Ikari NM, Peres SV, Lopes MAB, Lopes LM. Resultados perinatais e seguimento em longo prazo de tumores cardíacos fetais: estudo de coorte histórica de 30 anos. Arquivos Brasileiros de Cardiologia. 2024;121:e20220469.

21. Haycock GB, Schwartz GJ, Wisotsky DH. Geometric method for measuring body surface area: a height-weight formula validated in infants, children, and adults. The Journal of pediatrics. 1978;93:62–66.

22. Bois D. A formula to estimate the approximate surface area if height and weight be known. 1916. Nutrition. 1989;5:303.

23. Mitchell C, Rahko PS, Blauwet LA, Canaday B, Finstuen JA, Foster MC, Horton K, Ogunyankin KO, Palma RA, Velazquez EJ. Guidelines for performing a comprehensive transthoracic echocardiographic examination in adults: recommendations from the American Society of Echocardiography. Journal of the American Society of Echocardiography. 2019;32:1–64.

24. Lopez L, Colan S, Stylianou M, Granger S, Trachtenberg F, Frommelt P, Pearson G, Camarda J, Cnota J, Cohen M. Relationship of echocardiographic Z scores adjusted for body surface area to age, sex, race, and ethnicity: the pediatric heart network normal echocardiogram database. Circulation: Cardiovascular Imaging. 2017;10:e006979.

25. Pettersen MD, Du W, Skeens ME, Humes RA. Regression equations for calculation of z scores of cardiac structures in a large cohort of healthy infants, children, and adolescents: an echocardiographic study. Journal of the American Society of Echocardiography. 2008;21:922–934.

26. Hosmer Jr DW, Lemeshow S, Sturdivant RX. Applied logistic regression. John Wiley & Sons; 2013.

27. Chen X-Q, Wang Y-Y, Zhang M-N, Lu Q, Pang L-Y, Liu L-Y, Li Y-F, Zou L-P. Sirolimus can increase the disappearance rate of cardiac rhabdomyomas associated with tuberous sclerosis: a prospective cohort and self-controlled case series study. The Journal of Pediatrics. 2021;233:150–155. e154.

28. Davis PE, Filip-Dhima R, Sideridis G, Peters JM, Au KS, Northrup H, Bebin EM, Wu JY, Krueger D, Sahin M. Presentation and diagnosis of tuberous sclerosis complex in infants. Pediatrics. 2017;140.

29. Kingswood C, Bolton P, Crawford P, Harland C, Johnson SR, Sampson JR, Shepherd C, Spink J, Demuth D, Lucchese L. The clinical profile of tuberous sclerosis complex (TSC) in the United Kingdom: a retrospective cohort study in the Clinical Practice Research Datalink (CPRD). european journal of paediatric neurology. 2016;20:296–308.

30. Northrup H, Koenig MK, Pearson DA, Au KS. Tuberous sclerosis complex synonym: Bourneville disease. Gene. 2018;1:2.

31. Jeong A, Wong M. Systemic disease manifestations associated with epilepsy in tuberous sclerosis complex. Epilepsia. 2016;57:1443–1449.

32. Ng SY, Luk H-M, Hau EW, Cheng SS, Yu KP, Ho S, Mok MT, Lo IF. Genotype/phenotype correlation in 123 Chinese patients with Tuberous Sclerosis Complex. European Journal of Medical Genetics. 2022;65:104573.

33. Amin S, Lux A, Calder N, Laugharne M, Osborne J, O’callaghan F. Causes of mortality in individuals with tuberous sclerosis complex. Developmental medicine & child neurology. 2017;59:612–617.

34. Bosi G, Lintermans J, Pellegrino P, Svaluto-Moreolo G, Vliers A. The natural history of cardiac rhabdomyoma with and without tuberous sclerosis. Acta Paediatrica. 1996;85:928–931.

35. Nir A, Tajik AJ, Freeman WK, Seward JB, Offord KP, Edwards WD, Mair DD, Gomez MR. Tuberous sclerosis and cardiac rhabdomyoma. The American journal of cardiology. 1995;76:419–421.

36. Yıldırım S, Aypar E, Aydın B, Akyüz C, Aykan HH, Ertuğrul İ, Karagöz T, Alehan D. Cardiac rhabdomyomas: clinical progression, efficacy and safety of everolimus treatment. The Turkish Journal of Pediatrics. 2023;65:479–488.

